# Social problems and brain structure development following childhood mild traumatic brain injury

**DOI:** 10.1101/2022.09.20.22280146

**Authors:** Fanny Dégeilh, Tilmann von Soest, Lia Ferschmann, Joanne C. Beer, Malo Gaubert, Inga K. Koerte, Christian K. Tamnes

**Author notes:** **Corresponding author:** Correspondence concerning this article should be addressed to Fanny Dégeilh, IRISA / Inria Rennes, Empenn, Campus Universitaire de Beaulieu, 35042 Rennes cedex – France. Co-last author (Inga K. Koerte, Christian K. Tamnes).

## Abstract

Childhood mild traumatic brain injury (mTBI) is associated with elevated risk of developing social problems, which may be underpinned by changes in the structural developmental trajectory of the social brain, a network of cortical regions supporting social cognition and behavior. However, limited sample sizes and cross-sectional designs generally used in neuroimaging studies of pediatric TBI have prevented explorations of this hypothesis. This longitudinal retrospective study examined the development of parent-reported social problems and cortical thickness in social brain regions following childhood mTBI using data from the large population-based Adolescent Brain Cognitive Development (ABCD) Study. Two-group latent change score models revealed different developmental trajectories from ages 10 to 12 years in social problems between children with (n=345) and without (n=7,089) mTBI. Children with mTBI showed higher levels of social problems than controls at age 10. Then, social problems decreased over 2 years, but still remained higher than in controls in which they stayed stable. Both groups showed similar decreases in social brain cortical thickness between ages 10 and 12 years. Further studies providing detailed information on the injury mechanism and acute symptoms are needed to better understand individual differences in social impairment and brain development in pediatric TBI.

## 1. Introduction

Traumatic brain injury (TBI) is highly prevalent in children and can lead to long-term behavioral problems, including social problems such as aggressive behaviors and social cognitive deficits (Anderson et al., 2022; Catroppa et al., 2015; Dégeilh et al., 2018; Li & Liu, 2013; Ryan et al., 2021; Zamani et al., 2019). Childhood TBI (i.e., sustained between 0 to 10 years of age), even of mild severity (mTBI) which represent 90% of TBI cases (Zemek et al., 2013), is associated with worse outcomes than TBI sustained in late adolescence (Donders & Warschausky, 2007) and may profoundly impact the child’s quality of life and academic achievements (Anderson et al., 2009). While the consequences of TBI on children’s social functions have received increased attention over the past decade, the brain mechanisms underlying the social dysfunction following childhood TBI remain poorly understood (Ryan, Catroppa, Godfrey, et al., 2016).

Social cognitive functions emerge early in development, but also show a protracted development, continuing across childhood and adolescence, coinciding with the extended maturation of the ‘social brain’, an anatomically distributed set of cortical regions underlying social cognition and behavior (Blakemore, 2012; Frith & Frith, 2003; Mills et al., 2014). The social brain comprises the posterior superior temporal sulcus, the anterior temporal cortex, the medial prefrontal cortex (medial Brodmann Area 10), and the inferior parietal cortex (also referred as the temporoparietal junction; Frith and Frith, 2003; Blakemore, 2012; Mills et al., 2014). Cortical thickness decreases in these brain regions from childhood through late adolescence (Mills et al., 2014; Tamnes et al., 2017). Childhood TBI may disrupt the expected brain structural developmental trajectories of social brain regions, that could manifest as development of social problems.

Despite the heterogeneity of the brain injury profiles following TBI, common causes of injury involve similar mechanisms, and some brain regions are more vulnerable than others to injury, and therefore more commonly damaged (Bigler, Abildskov, et al., 2013). Particularly vulnerable brain regions include parts of the social brain network. Indeed, given the shape of the skull and how the brain is held in situ, focal contusions often occur in frontotemporal regions and lead to a frontotemporal distribution of cortical atrophy (Bigler, Abildskov, et al., 2013). Moreover, evidence from neuroimaging studies suggest an association between structural social brain alterations (i.e., brain lesion, reduced cortical volume or thickness in frontal, temporal, and parietal regions) and persistent social deficits following pediatric moderate-to-severe TBI (Bigler, Yeates, et al., 2013; Levan et al., 2015; Ryan, Catroppa, Beare, et al., 2016; Ryan, Catroppa, Godfrey, et al., 2016; Ryan, van Bijnen, et al., 2016; Zamani et al., 2019). Concerning mTBI, the very few studies conducted so far show conflicting results, with some reporting reduced gray matter volumes and cortical thickness in frontal, temporal and parietal regions following pediatric mTBI (Mayer et al., 2015; Urban et al., 2017), while others report no changes (King et al., 2019; Ryan, Catroppa, Beare, et al., 2016).

The interaction between injury physiopathology and brain maturation may underpin the long-term functional impact of pediatric TBI (King et al., 2019). Longitudinal studies of brain maturation following pediatric TBI are however scarce. The three available studies explored longitudinal changes in regional brain volumes and cortical thickness, and suggested deviation from the expected developmental pattern, particularly in frontal and temporal regions, following mTBI or moderate-to-severe TBI sustained in late childhood and adolescence (Dennis et al., 2017; Mayer et al., 2015; Wilde et al., 2012). However, the small sample sizes (all Ns < 21 TBI) and analytic methods not handling challenges associated with longitudinal data, such as partially missing data, limit the generalization of these findings. In addition, none of these studies included children who sustained a TBI at a younger age than 8 years.

The present study aimed to characterized longitudinal changes in social problems and cortical thickness in social brain regions following pediatric mTBI using data from the large Adolescent Brain Cognitive Development (ABCD) Study (Casey et al., 2018). We used an analytic method sensitive to inter-individual variability in intra-individual change, namely latent change score models (LCSM), to test for group differences in developmental changes in social problems and cortical thickness from ages 10.0 years to ages 11.9 years in 224 children with a history of childhood mTBI and 5,736 controls.

## 2. Methods

### 2.1. Study design

Data used in the current analyses was collected as part of the ABCD Study®. The ABCD study is a large 21-site longitudinal study of child development in the United States, which recruited 11,876 children aged 8.9 to 11.1 years between 2016 and 2018 and follow them annually (Barch et al., 2018; Garavan et al., 2018). All ABCD study procedures were approved by the local human research ethics committees. All families provided written informed consent for participation.

### 2.2. Measures

The current study used baseline, 1-year follow-up and 2-year follow-up demographic, physical health, mental health (Barch et al., 2018), and neuroimaging (Casey et al., 2018) data from ABCD 4.0 data release (http://dx.doi.org/10.15154/1527902). Only measures included in the current analyses are described here.

#### 2.2.1. Demographic Variables

Demographic variables, including child age at each visit, sex, and parental education (i.e., highest grade or level of school completed or highest degree received), were reported by parents via the demographic questionnaire (Barch et al., 2018).

#### 2.2.2. Brain injury history

The Modified Ohio State University TBI Screen-Short Version (Barch et al., 2018; Bogner et al., 2017; Corrigan & Bogner, 2007) is a parent-report questionnaire designed to measure lifetime history of TBI in children. Parents are asked to recall events related to the occurrence of injury to the head or neck (e.g., car accident, fall, being hit by someone or something). In case of positive response to any occurrence, follow-up questions are asked to determine the age of the injury and whether the injury was associated with loss of consciousness (LOC) or memory loss. At baseline, parents were asked about the child’s entire life (i.e., the first 10 years of life). At follow-ups, parents were asked to report on any new injury to the head or neck that may have occurred since the previous visit.

The summary score (‘tbi_ss_worst_overall’) was used to identify participants as mTBI or controls. The summary score rates injury severity on a five-point scale: (**1**) Improbable TBI (no TBI or TBI without LOC or memory loss), (**2**) Possible mild TBI (TBI without LOC, but with memory loss), (**3**) Mild TBI (TBI with LOC ≤ 30 minutes), (**4**) Moderate TBI (TBI with LOC between 30 minutes to 24 hours), and (**5**) Severe TBI (TBI with LOC ≥ 24 hours). Children with a moderate or severe TBI (score > 3) at baseline, who reported sustaining a TBI between baseline and 2-year follow-up visit, or which the score was missing were excluded from the analyses. Children with a score of 2 or 3 at baseline were included in the mTBI group. Children with a summary score of 1 <u>without</u> history of neck or head injury at baseline or at follow-ups were included in the control group.

#### 2.2.3. Social problems

Baseline and 2-year follow-up age-corrected t-scores from the social problem subscale on the Child Behavior Checklist-Parent form (CBCL; Achenbach and Rescorla, 2000) were used as measures of children’s social problems. CBCL is a parent report of emotional and behavioral problems in children and adolescents. Each item is rated on a three-point rating scale (0 = Not true; 1 = Somewhat or sometimes true; 2 = Very true or often true), and the social problems subscale includes 11 items.

#### 2.2.4. Social brain cortical thickness

Baseline and 2-year follow-up cortical thickness was computed on T1-weighted images by the ABCD group (Hagler et al., 2019) using FreeSurfer v5.3 (Fischl, 2012) within the Desikan-Killiany regions (Desikan et al., 2006). The following left and right hemisphere regions of interest (ROIs) corresponding to core region of the social brain network (Mills et al., 2014) were used in our analyses: medial orbitofrontal cortex, anterior temporal cortex (temporal pole), inferior parietal cortex, and banks of the superior temporal sulcus (Fig. 1).

**Figure 1.**
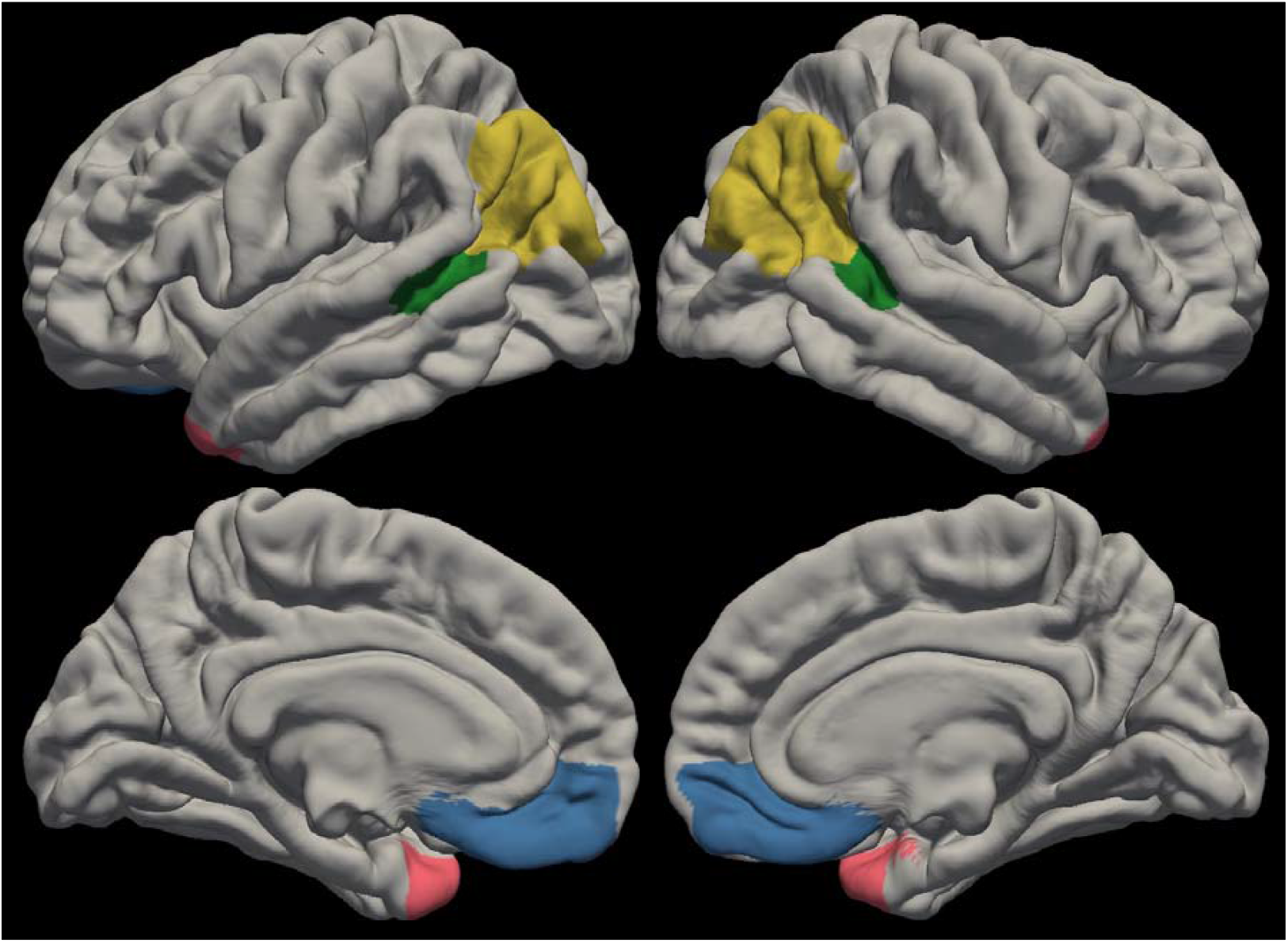
The social brain. Social brain regions of interest. medial orbitofrontal (blue); inferior parietal (yellow); superior temporal sulcus (green); anterior temporal cortex (temporal pole; pink)

Scanner effects (n scanners = 26) were corrected before statistical analyses using longitudinal ComBat (long-ComBat; Beer et al., 2020). Long-ComBat is an empirical Bayesian method for harmonizing longitudinal multi-scanner MRI data removing both additive and multiplicative scanner effects. ComBat is more robust to outliers in the case of small within-scanner sample sizes than other harmonization methods (Johnson et al., 2007). Long-ComBat model included explanatory variables child sex (0 = girl; 1 = boy) and parental education (ranging from 0 to 18), group, time x group interaction, and a subject-level random intercept. Fig. 2 shows residual boxplots by scanner before and after data harmonization using long-ComBat.

**Figure 2.**
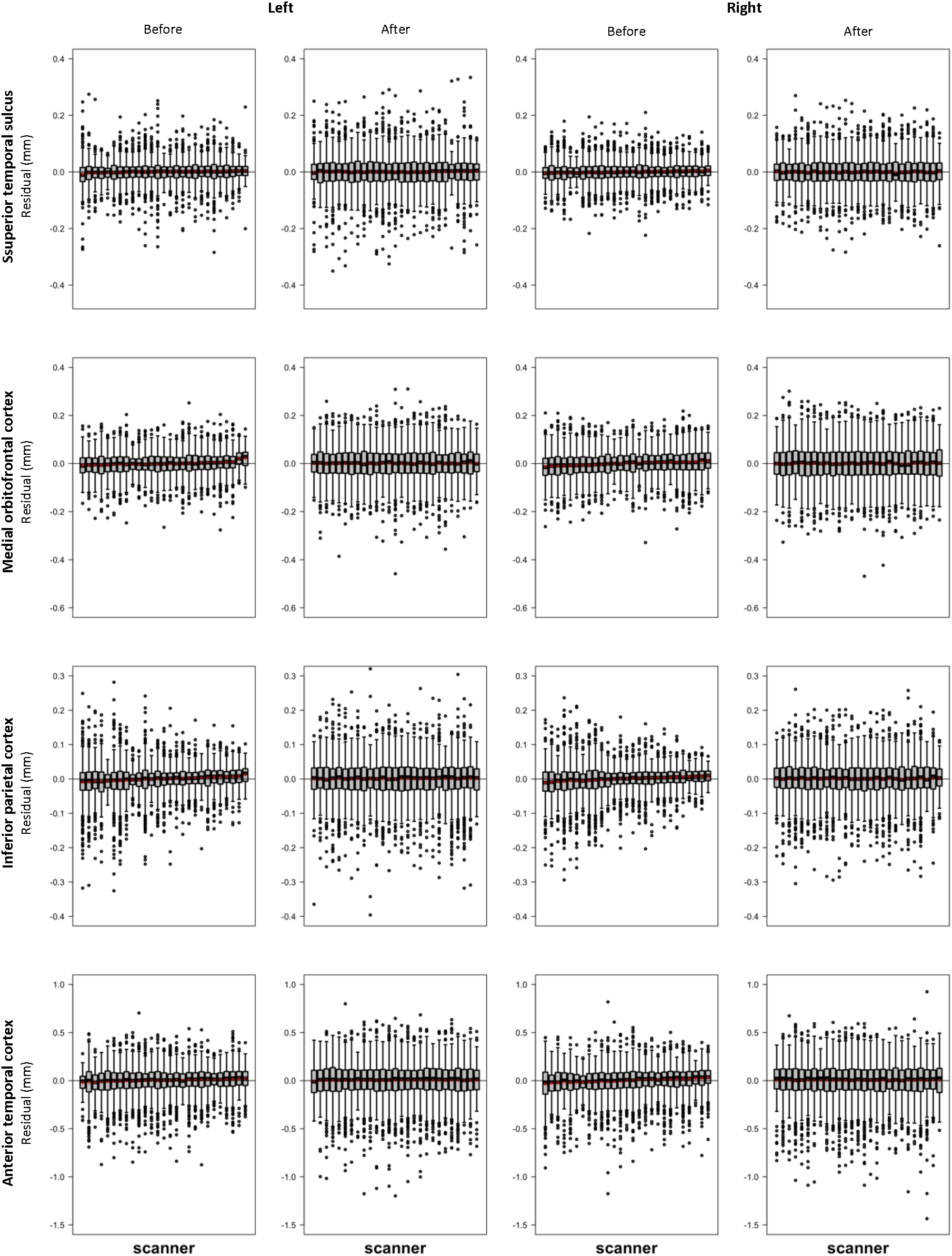
Distribution of cortical thickness residuals and means (red points) across scanner before and after harmonization with longitudinal ComBat for left and right social brain regions. Residuals are derived from linear mixed effects models including explanatory variables child sex (0 = girl; 1 = boy) and parental education (ranging from 0 to 18), group, time x group interaction, and a subject-level random intercept.

### 2.3. Statistical analyses

Group differences on demographic variables were tested using Chi-squared test (categorial data) and t-test (continuous data).

Multigroup LCSM (Kievit et al., 2018) were constructed with the lavaan 0.6-12 package (Rosseel, 2012) in R (version R4.1.2) to estimate latent change scores between baseline and follow-up for social problems and cortical thickness (for each ROI – Combat-corrected cortical thickness values) for all individuals (see Fig. 3). Group (mTBI versus controls) differences in four parameters of interest – the mean of the baseline score for behavior or cortical thickness, the mean of the latent change factor (reflecting rate of change over time), the variance of the baseline score (reflecting individual differences in scores at baseline) and the variance of the latent change factors (reflecting individual differences in rate of change) – were tested by means of equality constraints (Kievit et al., 2018). More specifically, each parameter of interest was individually constrained to be equal in the two groups and this constrained model was compared to a model where the parameter of interest was freely estimated using scaled Chi-squared difference tests (Satorra & Bentler, 2001). A significant difference between the two models reflects that the constrained parameter is significantly different between the two groups. Both the baseline measure and the latent change factor were controlled for child sex (0 = girl; 1 = boy) and parental education (ranging from 0 to 18).

**Figure 3:**
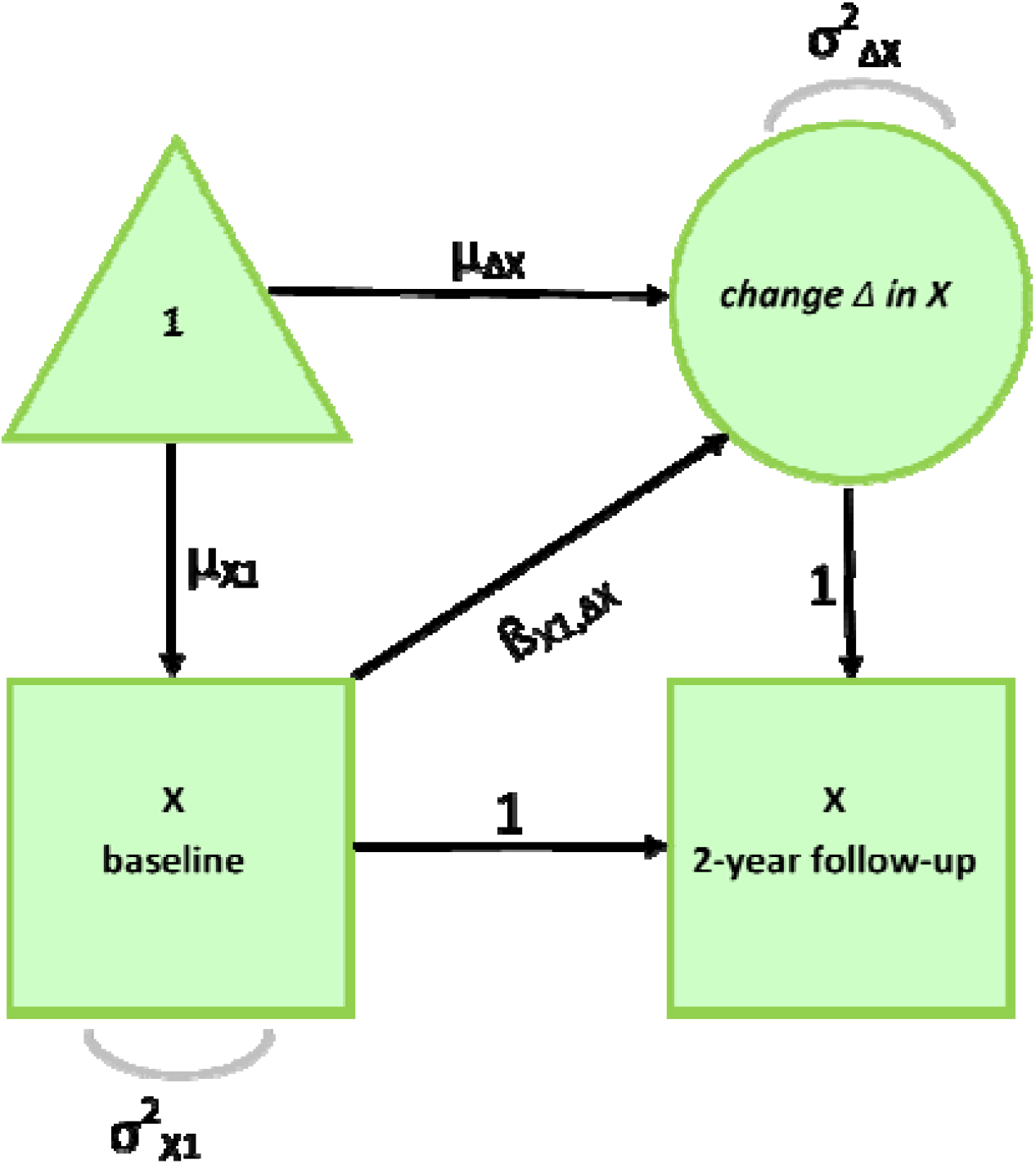
Latent Change Score Model. Social problems and cortical thickness (X) measured at baseline and at 2-year follow-up. Change (Δ) in X across time is modeled as latent variable. Sex and parental education were entered as covariates.

When the multigroup LCSM reveled significant results, a secondary univariate LCSM was constricted to the mTBI group to explore if the changes observed following mTBI were associated with age at injury.

## 3. Results

### 3.1. Participants final sample

From the 11,876 participants of ABCD data release 4.0, 4,318 were not eligible for the current study or met an exclusion criterion, 351 children were categorized as mTBI and 7,207 children were categorized as controls (Figure 4). Of these 7,558 eligible participants, 124 were excluded from the analyses because of the following criteria: Structural MRI data were missing at both timepoint for 79 controls and 2 mTBI, and did not pass the ABCD quality control (Hagler et al., 2019) for 15 controls. To reduce imbalance in scanners between the two groups, 15 controls were excluded because their MRI data were collected on a scanner used for acquiring less than two scans per group (Beer et al., 2020). One participant was mislabeled as mTBI, and 3 had repetitive head impact. Finally, parental education (used as covariate in the analyses) was missing for 9 controls. The final sample for our analyses thus included 345 children with mTBI and 7,089 controls (cf. Table 1 for demographic information and group comparison). The two groups were significantly different regarding sex and parental education; thus, these two variables were entered as covariates in the analyses.

**Figure 4:**
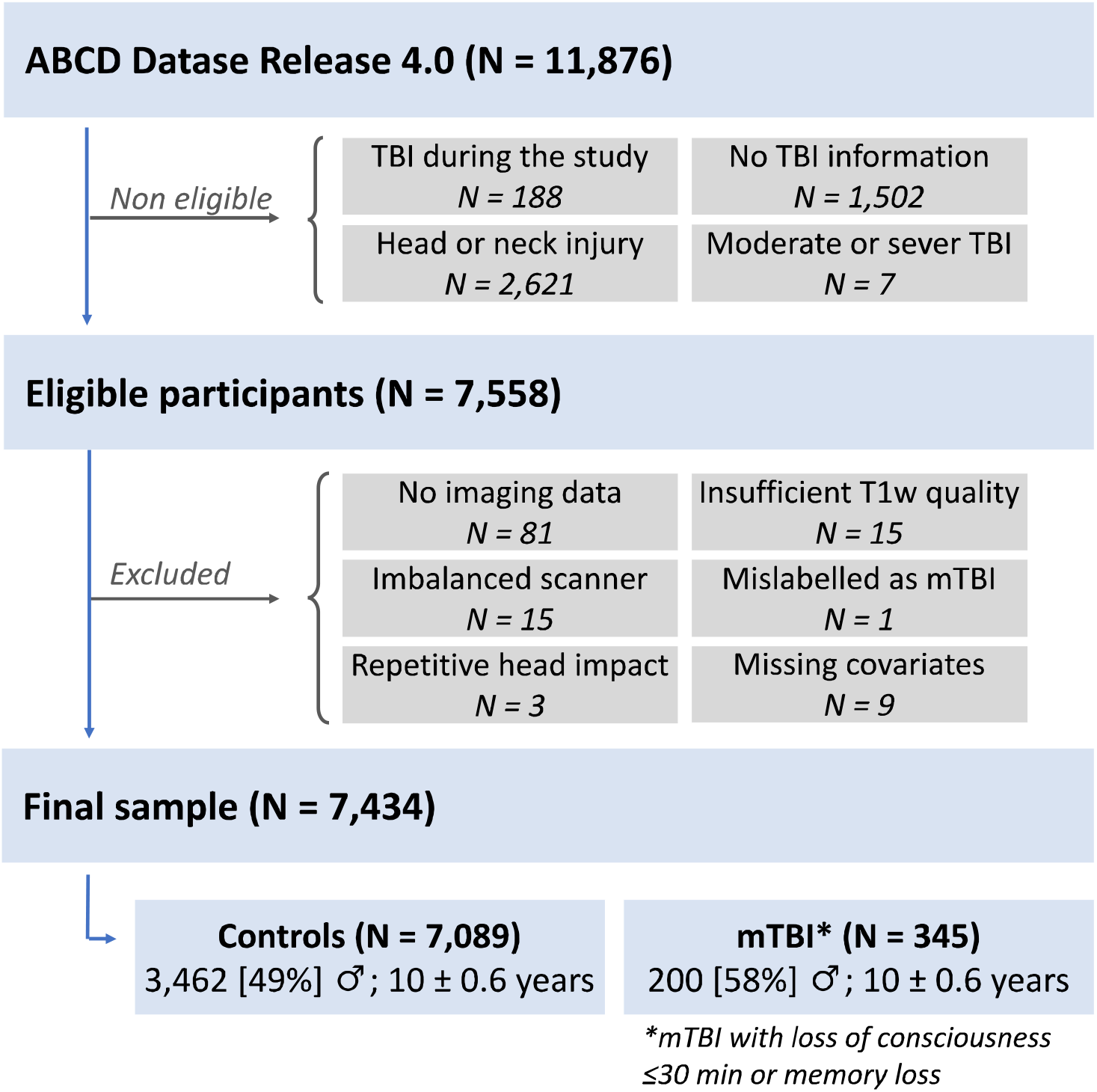
Participants selection

**Table 1:**
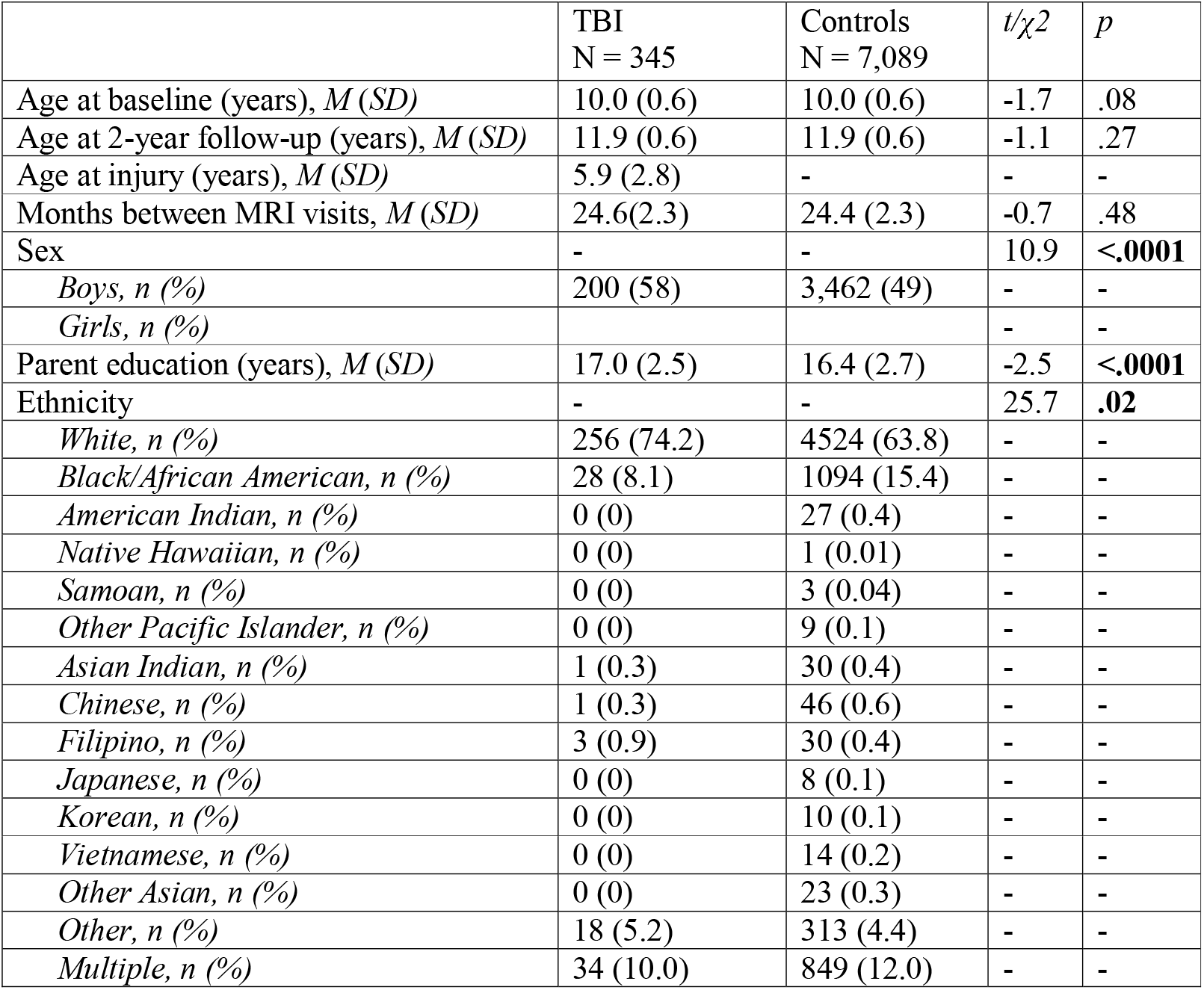
Demographic characteristics of the final sample

| | TBI<br>N = 345 | Controls<br>N = 7,089 | $t/\chi^2$ | $p$ |
| --- | --- | --- | --- | --- |
| Age at baseline (years), $M (SD)$ | 10.0 (0.6) | 10.0 (0.6) | -1.7 | .08 |
| Age at 2-year follow-up (years), $M (SD)$ | 11.9 (0.6) | 11.9 (0.6) | -1.1 | .27 |
| Age at injury (years), $M (SD)$ | 5.9 (2.8) | - | - | - |
| Months between MRI visits, $M (SD)$ | 24.6(2.3) | 24.4 (2.3) | -0.7 | .48 |
| Sex | - | - | 10.9 | **<.0001** |
| Boys, $n (%)$ | 200 (58) | 3,462 (49) | - | - |
| Girls, $n (%)$ | | | - | - |
| Parent education (years), $M (SD)$ | 17.0 (2.5) | 16.4 (2.7) | -2.5 | **<.0001** |
| Ethnicity | - | - | 25.7 | **.02** |
| White, $n (%)$ | 256 (74.2) | 4524 (63.8) | - | - |
| Black/African American, $n (%)$ | 28 (8.1) | 1094 (15.4) | - | - |
| American Indian, $n (%)$ | 0 (0) | 27 (0.4) | - | - |
| Native Hawaiian, $n (%)$ | 0 (0) | 1 (0.01) | - | - |
| Samoan, $n (%)$ | 0 (0) | 3 (0.04) | - | - |
| Other Pacific Islander, $n (%)$ | 0 (0) | 9 (0.1) | - | - |
| Asian Indian, $n (%)$ | 1 (0.3) | 30 (0.4) | - | - |
| Chinese, $n (%)$ | 1 (0.3) | 46 (0.6) | - | - |
| Filipino, $n (%)$ | 3 (0.9) | 30 (0.4) | - | - |
| Japanese, $n (%)$ | 0 (0) | 8 (0.1) | - | - |
| Korean, $n (%)$ | 0 (0) | 10 (0.1) | - | - |
| Vietnamese, $n (%)$ | 0 (0) | 14 (0.2) | - | - |
| Other Asian, $n (%)$ | 0 (0) | 23 (0.3) | - | - |
| Other, $n (%)$ | 18 (5.2) | 313 (4.4) | - | - |
| Multiple, $n (%)$ | 34 (10.0) | 849 (12.0) | - | - |

### 3.2. Social problems

The two-group LCSM without constraints across groups (i.e., all parameters freely estimated) fits the data well: χ^2^ (4) = 19.12, Comparative Fit Index (CFI) = 0.99, Turkey-Lewis Index (TLI) = 0.98, Root Mean Square Error of Approximation (RMSEA) = 0.032 [0.019 0.047]. Constraining the parameters of interest one by one to be equal across groups significantly reduced the model fit in all models (all *p*s ≤ .002) reflecting group differences in both means and variances of the baseline value of and change in social problems (Table 2).

**Table 2:** Social problems - Model fit comparisons

| | df | $\chi^2$ | $\chi^2\Delta$ | *p* |
| --- | --- | --- | --- | --- |
| All parameters freely estimated | 4 | 19.46 |  |  |
| Baseline means constrained across groups | 5 | 47.21 | 87.35 | <.0001 |
| Baseline variances constrained across groups | 5 | 215.02 | 85.78 | <.0001 |
| Latent change factor means constrained across groups | 5 | 27.24 | 9.93 | .002 |
| Latent change factor variance constrained across groups | 5 | 66.49 | 14.76 | .0001 |

Children with a history of mTBI showed higher levels of social problems at baseline (estimated mean = 56.4) than the control group (estimated mean = 54.4; Table 3). Moreover, social problems decreased faster in children with history of mTBI (estimated mean = -1.2, *p* = .007) than in controls in which social problem score stayed stable over time (estimated mean: -0.3, *p* = .33; Table 3). Welch two sample t-test revealed that social problems at 2-year follow-up remained significantly higher (t = -3.47, df = 282.13, *p* = .0006) in children with an history of mTBI (mean = 53.6 +/- 6.01) than in controls (mean = 52.4 +/- 4.40). In addition, the proportion of children with clinically significant social problems was higher in mTBI than in controls at both timepoint (Table 4).

**Table 3:** Social problems – Model estimates (all parameters freely estimated)

|  | **Controls** | **TBI** |
| --- | --- | --- |
| Baseline means | 54.41* | 56.38* |
| Baseline variances | 17.83* | 45.70* |
| Latent change factor means | -0.29 | -1.16* |
| Latent change factor variance | 15.65* | 26.93* |
| Baseline regressed on sex ($\beta$) | 0.72* | |
| Change regressed on sex ($\beta$) | -0.37* | |
| Baseline regressed on parental education ($\beta$) | -0.18* | |
| Change regressed on parental education ($\beta$) | 0.03 | |

\* *p*-value < .05.

**Table 4:** Proportion of children with borderline, clinical, and non-clinical social problems in each group and at each timepoint

| | **Borderline**<br>**([65-69])** | **Clinical**<br>**(>=70)** | **Non-**<br>**Clinical**<br>**(<=64)** | **<NA>** | $\chi^2$ | *p* |
| --- | --- | --- | --- | --- | --- | --- |
| **Baseline** |  |  |  |  |  |  |
| TBI | 17 (4.93) | 19 (5.51) | 309 (89.57) | 0 (0) | 66.03 | <.0001 |
| Controls | 124 (1.75) | 81 (1.14) | 6,883 (97.09) | 1 (0.01) |  |  |
| **2-year follow-up** |  |  |  |  |  |  |
| TBI | 9 (2.61) | 10 (2.90) | 250 (72.46) | 76 (22.18) | 18.62 | <.0001 |
| Controls | 134 (1.89) | 54 (0.76) | 5,329 (75.17) | 1,572 (22.03) |  |  |

Children with mTBI showed greater individual difference at baseline (variance = 45.7) and in change (variance = 26.9) than the controls (baseline variance = 17.8, change variance = 15.6; Table 3). Of note, the results were similar when covariates (sex and parental education) were not included in the model.

Sex had significant association with both baseline levels and change in social problems (all *p*s <.0001; Table 3). Boys showed higher baseline scores and faster decrease over time in social problems than girls. Parent education had a negative significant association with baseline levels in social problems (all *p* <.0001; Table 3).

Secondary univariate LCSM in the mTBI group revealed no association between the age at injury and the means of the baseline value (p = .20) of and change in (p = .63) social problems.

### 3.3. Social brain cortical thickness

For all ROIs (medial orbitofrontal cortex, anterior temporal cortex, inferior parietal cortex, banks of the superior temporal sulcus), the two-group latent change score model without constraints across groups fit the data well: CFI were all equal to 1.00, TLI were between 0.99 and 1.00, and RMSEA were between 0.000 and 0.016 (Table 5). For all ROIs, but the left medial orbitofrontal cortex, constraining the parameters of interest to be equal between groups did not significantly reduce the model fit (all *p*s ≥ .12), reflecting no group difference on means and variances of the baseline value and change of the social brain cortical thickness (Table 6). Concerning the medial orbitofrontal cortex, constraining the variances of the baseline parameter to be equal across groups significantly reduced the model fit (all *p* = .03), reflecting group differences in the variances of change in social problems (Table 6).

**Table 5:** Social brain cortical thickness – Model fit (all parameters freely estimated)

| | $\chi^2$ (df = 4) | CFI | TLI | RMSEA [90CI] |
| --- | --- | --- | --- | --- |
| Left banks of the superior temporal sulcus | 3.82 | 1.00 | 1.00 | 0.000 [0.000 0.019] |
| Right banks of the superior temporal sulcus | 1.86 | 1.00 | 1.00 | 0.004 [0.000 0.0126] |
| Left medial orbitofrontal cortex | 5.27 | 1.00 | 0.99 | 0.016 [0.000 0.034] |
| Right medial orbitofrontal cortex | 4.31 | 1.00 | 1.00 | 0.007 [0.000 0.027] |
| Left inferior parietal cortex | 4.15 | 1.00 | 1.00 | 0.000 [0.000 0.019] |
| Right inferior parietal cortex | 2.43 | 1.00 | 1.00 | 0.011 [0.000 0.029] |
| Left anterior temporal cortex | 4.92 | 1.00 | 0.99 | 0.015 [0.000 0.033] |
| Right anterior temporal cortex | 3.99 | 1.00 | 0.99 | 0.011 [0.000 0.030] |

*Abbreviations: CFI, Comparative Fit Index; RMSEA, Root Mean Square Error of*
*Approximation; TLI, Turkey-Lewis Index; 90CI, 90% confidence interval.*

**Table 6:** Social brain cortical thickness - Models fit comparisons

| | Df | $\chi^2$ | $\chi^2\Delta$ | p |
| --- | --- | --- | --- | --- |
| **Left banks of the superior temporal sulcus** |  |  |  |  |
| All parameters freely estimated | 4 | 2.51 | - | - |
| Baseline means constrained across groups | 5 | 3.35 | 0.86 | .48 |
| Baseline variances constrained across groups | 5 | 2.62 | 0.11 | .34 |
| Latent change factor means constrained across groups | 5 | 3.31 | 0.08 | .37 |
| Latent change factor variance constrained across groups | 5 | 3.19 | 0.26 | .61 |
| **Right banks of the superior temporal sulcus** |  |  |  |  |
| All parameters freely estimated | 4 | 4.14 | - | - |
| Baseline means constrained across groups | 5 | 5.10 | 0.98 | .32 |
| Baseline variances constrained across groups | 5 | 2.12 | 0.19 | .66 |
| Latent change factor means constrained across groups | 5 | 4.23 | 0.09 | .76 |
| Latent change factor variance constrained across groups | 5 | 8.18 | 2.35 | .12 |
| **Left medial orbitofrontal cortex** |  |  |  |  |
| All parameters freely estimated | 4 | 7.13 | - | - |
| Baseline means constrained across groups | 5 | 7.21 | 0.08 | .78 |
| Baseline variances constrained across groups | 5 | 7.16 | 0.03 | .87 |
| Latent change factor means constrained across groups | 5 | 7.23 | 0.09 | .76 |
| Latent change factor variance constrained across groups | 5 | **12.76** | **4.56** | **.03** |
| **Right medial orbitofrontal cortex** |  |  |  |  |
| All parameters freely estimated | 4 | 4.47 | - | - |
| Baseline means constrained across groups | 5 | 4.96 | 0.49 | .48 |
| Baseline variances constrained across groups | 5 | 4.48 | 0.007 | .93 |
| Latent change factor means constrained across groups | 5 | 4.52 | 0.04 | .83 |
| Latent change factor variance constrained across groups | 5 | 4.83 | 0.31 | .57 |
| **Left inferior parietal cortex** |  |  |  |  |
| All parameters freely estimated | 4 | 1.65 | - | - |
| Baseline means constrained across groups | 5 | 2.74 | 1.06 | .30 |
| Baseline variances constrained across groups | 5 | 3.62.83 | 0.87 | .35 |
| Latent change factor means constrained across groups | 5 | 1.68 | 0.02 | .87 |
| Latent change factor variance constrained across groups | 5 | 3.06 | 0.48 | .49 |
| **Right inferior parietal cortex** |  |  |  |  |
| All parameters freely estimated | 4 | 5.57 | - | - |
| Baseline means constrained across groups | 5 | 5.62 | 0.05 | .82 |
| Baseline variances constrained across groups | 5 | 7.57 | 1.54 | .21 |
| Latent change factor means constrained across groups | 5 | 6.20 | 0.68 | .41 |
| Latent change factor variance constrained across groups | 5 | 6.28 | 0.33 | .56 |

| **Left anterior temporal cortex** |  |  |  |  |
| --- | --- | --- | --- | --- |
| All parameters freely estimated | 4 | 6.29 | - | - |
| Baseline means constrained across groups | 5 | 6.34 | 0.06 | .81 |
| Baseline variances constrained across groups | 5 | 7.46 | 0.80 | .53 |
| Latent change factor means constrained across groups | 5 | 6.83 | 0.58 | .45 |
| Latent change factor variance constrained across groups | 5 | 6.40 | 0.03 | .86 |
| **Right anterior temporal cortex** |  |  |  |  |
| All parameters freely estimated | 4 | 5.04 | - | - |
| Baseline means constrained across groups | 5 | 5.05 | 0.007 | .93 |
| Baseline variances constrained across groups | 5 | 5.75 | 0.25 | .61 |
| Latent change factor means constrained across groups | 5 | 5.05 | 0.006 | .94 |
| Latent change factor variance constrained across groups | 5 | 5.53 | 0.25 | .62 |

In both group and for all ROIs, except for the left and right anterior temporal cortex, thickness significantly decreased by 0.06 to 0.08 mm over the two years of the study period (Table 7). Sex was significantly associated with cortical thickness at baseline in the left and right banks of the superior temporal sulcus, inferior parietal cortex, and anterior temporal cortex, with boys showing thicker cortex than girls (Table 7). There were also significant sex differences in the rate of change of cortical thickness bilaterally in the banks of the superior temporal sulcus, the inferior parietal cortex, and the medial orbitofrontal cortex, with a faster decrease in boys than in girls. Parental education was significantly associated with cortical thickness in all ROIs at baseline, with higher parental education associated with thicker cortex (Table 7). Parental education was also significantly associated with the rate of change of cortical thickness in the left and right banks of the superior temporal sulcus and the left medial orbitofrontal cortex, with higher parental education associated with lower decrease.

**Table 7:** Social brain cortical thickness – Model estimates (all parameters freely estimated)

|  | Left | Right |
| --- | --- | --- |
| **Banks of the superior temporal sulcus** |  |  |
| Baseline means | **2.68\*** | **2.78\*** |
| Baseline variances | **0.02\*** | **0.03\*** |
| Latent change factor means | **-0.07\*** | **-0.06\*** |
| Latent change factor variance | **0.01\*** | **0.01\*** |
| Baseline regressed on sex ($\beta$) | **-0.02\*** | **-0.01\*** |
| Change regressed on sex ($\beta$) | **0.01\*** | **0.02\*** |
| Baseline regressed on parental education ($\beta$) | **0.004\*** | **0.005\*** |
| Change regressed on parental education ($\beta$) | **0.001\*** | **0.001\*** |
| **Medial orbitofrontal cortex** |  |  |
| Baseline means | **2.89\*** | **2.83\*** |
| Baseline variances | **0.02\*** | **0.02\*** |
| Latent change factor means | **-0.08\*** | **-0.06\*** |
| Latent change factor variance | **0.01\*<sup>b</sup>** | **0.02\*** |
| Baseline regressed on sex ($\beta$) | 0.003 | -0.005 |
| Change regressed on sex ($\beta$) | **0.01\*** | **0.01\*** |
| Baseline regressed on parental education ($\beta$) | **0.002\*** | **0.002\*** |
| Change regressed on parental education ($\beta$) | **0.002\*** | 0.001 |

\* $p$ -value < .05. <sup>b</sup> TBI (0.011) significantly lower than controls (0.014).

|  | Left | Right |
| --- | --- | --- |
| **Inferior parietal cortex** |  |  |
| Baseline means | **2.70\*** | **2.73\*** |
| Baseline variances | **0.01\*** | **0.01\*** |
| Latent change factor means | **-0.06\*** | **-0.06\*** |
| Latent change factor variance | **0.01\*** | **0.01\*** |
| Baseline regressed on sex ($\beta$) | **-0.02\*** | **-0.02\*** |
| Change regressed on sex ($\beta$) | **0.02\*** | **0.02\*** |
| Baseline regressed on parental education ($\beta$) | **0.003\*** | **0.002\*** |
| Change regressed on parental education ($\beta$) | 0.001 | 0.00 |
| **Anterior temporal cortex** |  |  |
| Baseline means | **3.57\*** | **3.71\*** |
| Baseline variances | **0.10\*** | **0.09\*** |
| Latent change factor means | -0.03 | -0.03 |
| Latent change factor variance | **0.10\*** | **0.10\*** |
| Baseline regressed on sex ($\beta$) | **-0.02\*** | **-0.02\*** |
| Change regressed on sex ($\beta$) | -0.01 | 0.01 |
| Baseline regressed on parental education ($\beta$) | **0.01\*** | **0.005\*** |
| Change regressed on parental education ($\beta$) | 0.001 | 0.002 |

Of note, the results were similar when covariates were not entered in the model, and the analyses were conducted on the raw (not harmonized for the scanners) MRI data.

## 4. Discussion

This study examined the longitudinal development of social problems and cortical thickness in social brain regions following childhood mTBI. Using a large sample with retrospective information on history of TBI and multigroup latent change score models, an analytic method sensitive to longitudinal change at both individual and group level, we found different developmental changes in social problems between children with and without history of mTBI. Children with mTBI showed higher levels of social problems than children without mTBI at age 10. Then, social problems decreased over 2 years but remained higher in children with mTBI than in controls in which they stayed stable. No group difference in the trajectory of the social brain cortical thickness was observed. Cortical thickness significantly decreased between age 10 and 12 years in both groups, except for the anterior temporal cortex where the thickness remained stable over the 2 years.

The present behavioral findings are consistent with previous studies showing persistent social impairments in children with TBI compared to their non-injured peers (Anderson et al., 2022; Catroppa et al., 2015; Dégeilh et al., 2018; Li & Liu, 2013; Ryan et al., 2021; Zamani et al., 2019). Here, we further observed that mTBI during childhood was associated with higher social problems, with a decrease over time that may suggest a slow recovery trajectory. Additional follow-up of the ABCD cohort will allow to test this hypothesis and explore whether or not adolescents with mTBI reach a similar level as their non-injured peers by the end of the adolescence.

While it is believed that the brain development and integrity may underpin social impairments developing following a pediatric TBI (Beauchamp & Anderson, 2010), no significant difference in the developmental trajectory of the cortical thickness in the social brain was observed in the current study. This result echoes recent findings by Anderson and colleagues (Anderson et al., 2022) showing no difference in gray matter volume of large-scale brain networks (i.e., Central Executive Network, Default Mode Network, Mentalizing Network, Mirror Neuron Empathy Network, and Salience Network) 3 to 8 weeks post-injury between TBI children with different profiles of social impairment recovery trajectory. Future studies should explore if changes in social brain function or other brain structure metrics may underly the changes in social problems after pediatric TBI.

In both groups, cortical thickness significantly decreased over the two years of the study in all the regions of the social brain but the anterior temporal cortex. This developmental trajectory of the social brain structure is consistent with what has been observed in longitudinal studies showing that the cerebral cortex undergoes widespread and regionally variable nonlinear decreases in thickness across childhood and adolescence (Mills et al., 2014; Tamnes et al., 2017). Concerning the social brain regions, cortical thickness in the posterior superior temporal sulcus, the medial prefrontal cortex, and the inferior parietal cortex has in previous studies consistently been found to decreased with age, while the findings for the anterior temporal cortex are less consistent, with both increase and decrease observed.

The present study was strengthened by its large sample of children who sustained a mTBI (n = 225, against n < 21 in published neuroimaging studies) between ages 0 to 9 years old (none of the published neuroimaging studies included children who sustained a mTBI before aged of 8 years old), and its longitudinal design. We applied a longitudinal analytic method well-suited to model individual differences in change across two time points and to handle missing data by means of full information maximum likelihood estimation. Furthermore, special attention was paid to scanner effects correction using a suitable method for longitudinal data.

Nonetheless, the current results must be interpreted in the context of some study limitations. Most notably, the assessment of TBI status was based on parent retrospective report and thus reflects parents’ memory of their child lifetime history of TBI, which is subject to recall bias in parental response to the questionnaire. In addition, the Modified Ohio State University TBI Screen-Short Version provides only an indirect index of child TBI history with limited information on mechanisms of injury and acute symptoms. Furthermore, the current mTBI definition (from Modified Ohio State University TBI Screen) is based on LOC or or memory loss duration allowing to identify only children having experienced more severe mTBI (complicated mTBI) and does not allow to identify children having experienced head or neck injury (excluded from the current analyses) in the absence of alterations in mental status for which acute symptoms is need to conclude on the presence or absence of a mTBI diagnose. Finally, given the absence of social problems measures prior to TBI, the possibility that higher social problems were already present in children prior to TBI cannot be excluded. Prospective longitudinal studies with more detailed information on the mechanisms and type of injury, acute symptoms and brain lesion, and objective assessment of the injury severity (i.e., Glasgow coma scale, symptoms measurement) providing a detailed picture of the pediatric TBI heterogeneity, are needed to better understand the neural mechanisms underpinning social impairment after a pediatric TBI.

## Data Availability

he data that support the findings are available from the NIMH Data Archive (NDA). The ABCD data used in this report came from the ABCD 4.0 data release [NIMH Data Archive Digital Object Identifier (DOI): http://dx.doi.org/10.15154/1527902].

## Data availability

The data that support the findings are available from the NIMH Data Archive (NDA). The ABCD data used in this report came from the ABCD 4.0 data release [NIMH Data Archive Digital Object Identifier (DOI): http://dx.doi.org/10.15154/1527902].

## Code availability

The code that supports the findings of this study is available as supplementary material.

## ABCD Study

Data used in the preparation of this article were obtained from the Adolescent Brain Cognitive Development^SM^ (ABCD) Study (https://abcdstudy.org), held in the NIMH Data Archive (NDA). This is a multisite, longitudinal study designed to recruit more than 10,000 children age 9-10 and follow them over 10 years into early adulthood. The ABCD Study® is supported by the National Institutes of Health and additional federal partners under award numbers U01DA041048, U01DA050989, U01DA051016, U01DA041022, U01DA051018, U01DA051037, U01DA050987, U01DA041174, U01DA041106, U01DA041117, U01DA041028, U01DA041134, U01DA050988, U01DA051039, U01DA041156, U01DA041025, U01DA041120, U01DA051038, U01DA041148, U01DA041093, U01DA041089, U24DA041123, U24DA041147. A full list of supporters is available at https://abcdstudy.org/federal-partners.html. A listing of participating sites and a complete listing of the study investigators can be found at https://abcdstudy.org/consortium_members/. ABCD consortium investigators designed and implemented the study and/or provided data but did not necessarily participate in the analysis or writing of this report. This manuscript reflects the views of the authors and may not reflect the opinions or views of the NIH or ABCD consortium investigators.

## Authors Contributions

FD conceptualized the study, conducted the analyses and drafted the original manuscript. TvS contributed to defining the statistical models, conducting the analyses and writing the manuscript. LF contributed to conducting the analyses and commented on the manuscript. JB contributed the data processing and commented on the manuscript. MG organized the data collection and commented on the manuscript. IKK contributed to conceptualizing the study, provided access to the data, and commented on the manuscript. CKT contributed to conceptualizing the study and writing the manuscript, and supervised the project. All authors contributed to the editing of the paper.

